# Unbiased estimation of the population-level motor module

**DOI:** 10.1101/2023.06.25.23291878

**Authors:** Yusuke Matsui, Kohei Uno, Ippei Nojima

**Affiliations:** Biomedical and Health Informatics Unit, Department of Integrated Health Science, Nagoya University Graduate School of Medicine, 461-8673 Nagoya, Aichi, Japan; Institute for Glyco-core Research (iGCORE), Nagoya University, 461-8673 Nagoya, Aichi, Japan; Graduate School of Medical Sciences, Department of Physical Therapy, Nagoya City University

**Keywords:** Motor module, computational approach, muscle activity, functional data analysis, estimation, model

## Abstract

Motor module is a functional neurophysiological command for muscle coordination. In clinical settings, population-level characterization and comparison of motor modules are necessary to evaluate pathophysiological mechanisms and intervention effects. Previous studies have estimated individual motor modules and then compared them, but the validity of capturing the distribution of the latent population has not been fully understood. Our study aimed to address this issue by investigating the accuracy of estimating the population mean of motor modules. Through simulation experiments, we found that previous individual-based approach did not converge regardless of sample size and was vulnerable to noise. We developed an unbiased estimation algorithm using the framework of functional data analysis, which significantly improved estimation accuracy. Our findings highlight statistical challenges for motor module analysis and suggest the need for further research on new computational algorithms using large-scale clinical data.

**Graphical Abstract:** 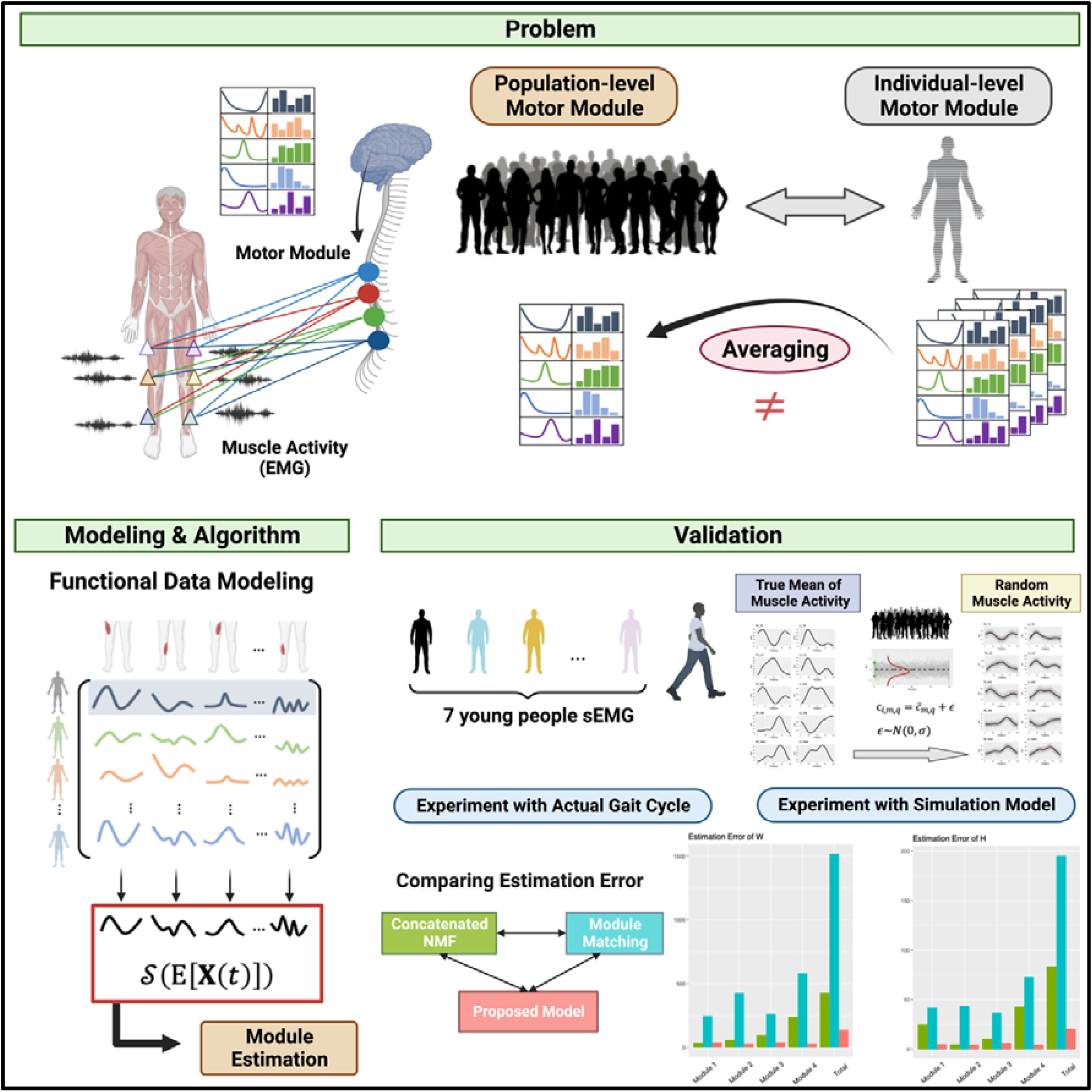

## Introduction

Motor control is a complex process that involves coordinated commands for thousands of motor units in hundreds of skeletal muscles, generated by the central nervous system in real-time to produce movements. To help explain this complexity, the concept of a motor module or muscle synergy has been proposed^1–6^. A motor module is a set of functional neurophysiological commands that drive muscle coordination^1,7^. Currently, motor modules are characterized via dimension-reduction techniques such as non-negative matrix factorization (NMF) ^8–10^(**Figure 1A**). However, NMF only finds two matrices that minimize residual errors for each dataset, and the resulting values have no biological meaning^11,12^. This makes it challenging to compare or summarize motor modules across multiple subjects (**Figure 1C**). Previous computational methodological study has mainly focused on developing sophisticated models for individual motor modules and improving their estimation reproducibility^13–15^. However, to gain a better understanding of the generality of motor modules, it is also crucial to establish their structure at the population level. Unfortunately, however, statistical and computational methods for this purpose have not yet been fully elucidated.

**Figure 1.**
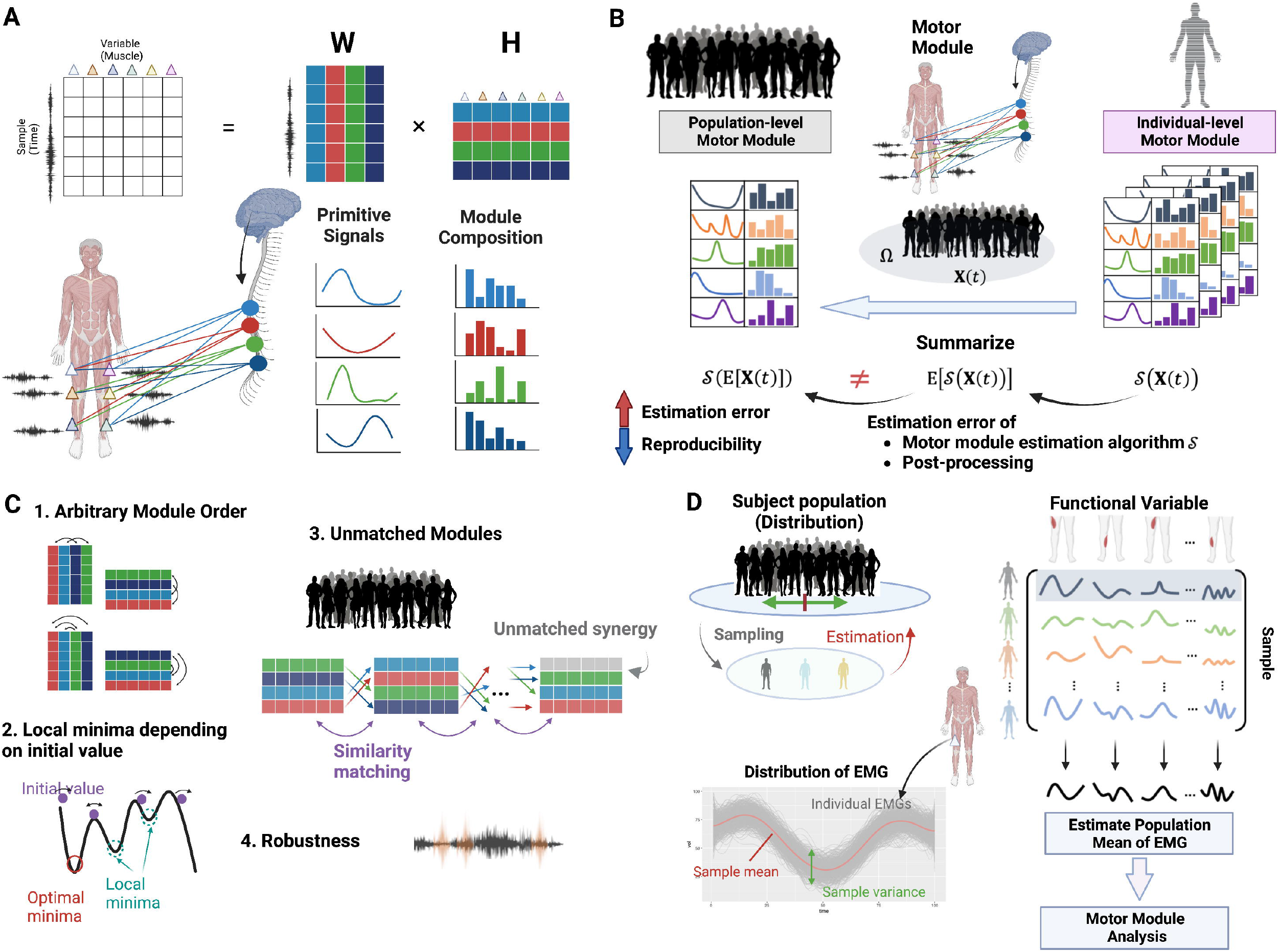
Conceptual view of this study. **A) Description of the general motor module estimation algorithm**. Multichannel Electromyography (EMG) data is given by a time × muscle matrix. Motor module analysis is a problem of unsupervised low-dimensional structural learning based on muscle activity data from multiple sites. Non-negative matrix factorization (NMF) is widely used as one of them. Each estimated dimension is considered to reflect the activity of the central nervous system, where **W** is called the primitive signal and is considered to be the input signal of the central nervous system, and **H** is called synergy and is considered to represent a potentially cooperating muscle group, or motor module. **B) Population-level motor module and individual-level module**. When conducting a multi-subject motor module analysis, there are two approaches depending on the purpose. The first is population-level motor module analysis, which is intended for characterization and inference at the population level and for comparison among populations. The second is individual-level motor module analysis, the main objectives of which are individual-level characterization and inter-individual comparisons. Our goal is to estimate population-level motor module expectations in an unbiased manner. We show that conventional methods are not consistent with true expectations where they estimate the expectation by aggregating individual-level motor modules. **C) Problems with conventional methods**. The main causes are 1) the arbitrariness of the solution due to the unsupervised learning of NMF, 2) local minima problem with an initial value, 3) errors associated with integrating the results of motor module analysis from different subjects, and 4) instability of the numerical solution of the NMF algorithm. **D) Proposed method**. We introduce the framework of functional data analysis. Assuming a stochastic population of muscle activity, the unbiased motor module expectation is estimated based on the expectation of muscle activity.

**Figure 2.**
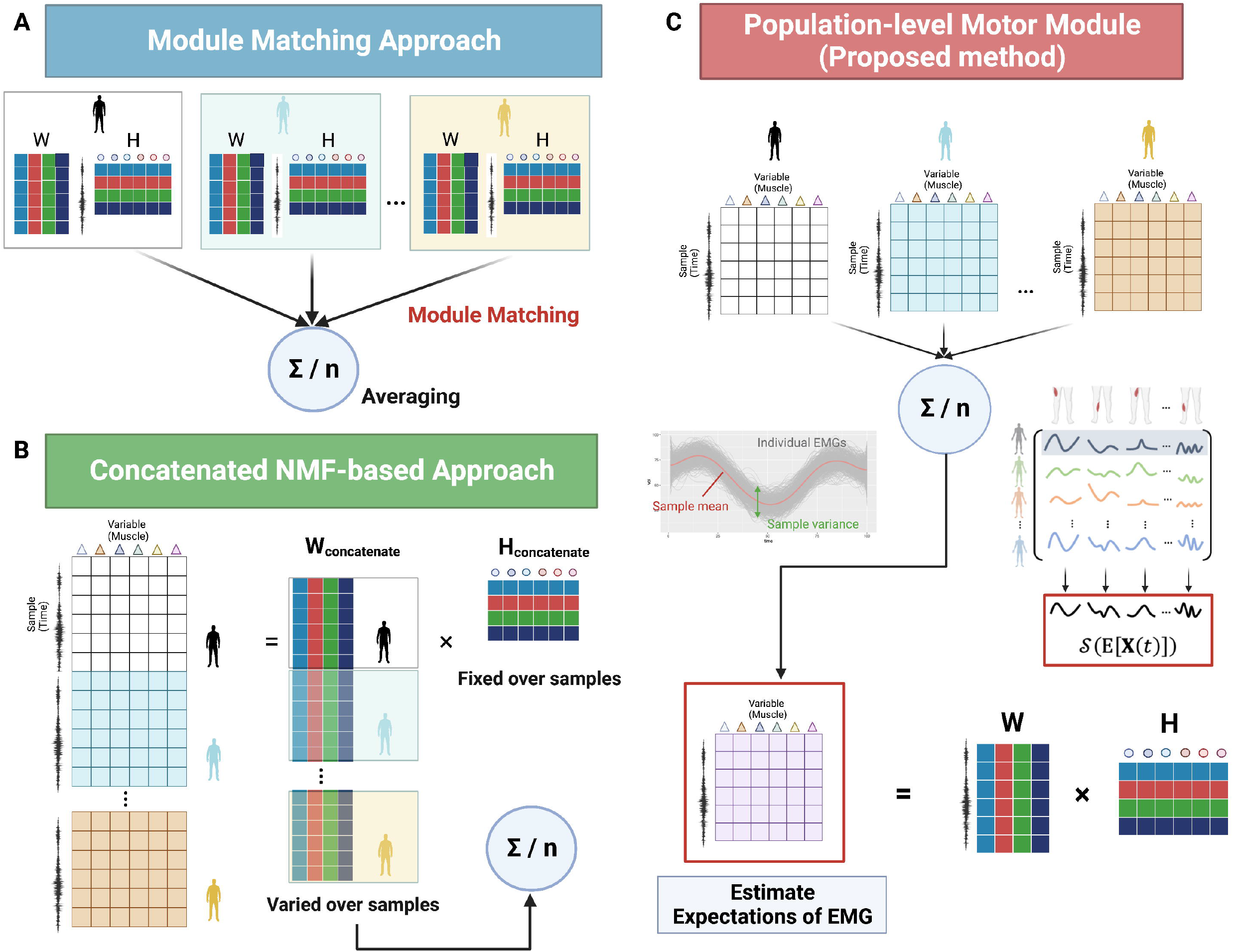
Comparison of algorithms between existing and proposed methods. **A) Module matching approach**. Multichannel Electromyography (EMG) data given in a time × muscle matrix for each individual was used to estimate the motor module using NMF, and then they are matched and averaged across subjects. **B) Concatenated NMF-based approach**. The NMF is applied by concatenating the EMG matrices across subjects in the row direction. Since the columns of the muscle activity matrix are fixed, the estimated synergy **H** approximates the average across subjects, and the primitive signal **W** conditioned by the common synergy **H** is estimated for each subject. **C) Proposed method**. First, expectations of muscle activity are estimated from the EMG matrices of multiple subjects, and NMF is applied to the obtained expected values of muscle activity. Since the expected values of EMG reflect the population structure, the obtained motor modules also reflect the population structure.

To address these challenges, two main approaches have been emerged. One approach involves module matching between subjects^16–20^, where individual motor modules are estimated and then matched to each other. However, this approach is not always guaranteed to give a one-to-one solution, and the repeated application of NMF individually can contaminate motor module estimates by local solutions due to initial value dependence. Another approach is joint learning called concatenated NMF (cNMF), where subjects’ data are combined into one pseudo-data as input^11,21,22^.This approach can help to avoid some of the problems with module matching, but it can also be numerically unstable and lead to convergence issues as the matrix size increases with the number of subjects. Overall, developing reliable statistical and computational methods for characterizing motor modules at the population level is an ongoing challenge, but it is crucial to advancing our understanding of motor control and its organization across individuals. To address these issues, this study presents an unbiased estimation algorithm for motor modules at the population level using NMF and Functional Data Analysis (FDA) ^23,24^ (**Figure 1D**). Our approach accounts for population variability and reduces statistical errors in motor module expectation estimation, particularly in larger sample sizes where other methods fail to converge.

## Results

### Population-level motor module estimation

In this section, we statistically formulated the population-level motor module borrowing the concept of FDA^23,24^. We decompose the variability in muscle activity into two levels: within-individual variability and between-individual variability. Within-individual variability accounts for variations within a single subject, such as trial-to-trial variation in the gait cycle, which can be reduced through preprocessing steps like filtering and temporal alignment. In contrast, between-individual variability arises from differences between subjects. We use the FDA to model this variability, taking into account the population characteristics. In the discussion that follows, individual muscle activity data shall be subjected to appropriate preprocessing, such as filtering, outlier removal, temporal alignment, and summarization, for inter-subject comparison.

To describe between-subjects variability in muscle activity, individual muscle activity patterns are described by functional random variables. Let *X*_*m*_(*t*) be a random functional variable, where the activity of skeletal muscle *m* at *t* time is represented by the function. The muscle activity *x*_*i,m*_(*t*) of subject *i* that we observe can be viewed as the observed values sampled from the functional population Ω. The observed muscle activity of individual subject can be denoted with the population mean

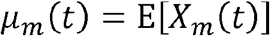

and the error term *ϵ*_*m*_(*t*) as

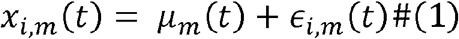

where *ϵ*_*m*_(*t*) is parameterized by E[*ϵ*_*m*_(*t*)] = 0 and Var[*ϵ*_*m*_(*t*)] = *σ*, representing population-level variability. The typical distribution that *X*_*m*_(*t*) follows is the normal distribution, but if non-negative constraints are considered, a non-negative exponential family of distribution (such as the Poisson distribution) can also be assumed^25^. In practice, to deal with digitized discrete data, time *t* is discretized as a vector *t* with length *T*:

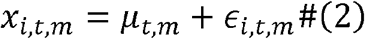

Our proposed population-level motor module can be estimated using *μ*_*t,m*_. The population mean of muscle activity, expressed in matrix form with *μ*_*t,m*_ as an element, is

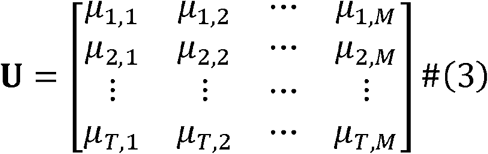

and motor module can be estimated by dimension reduction algorithm. In case of NMF, the mean muscle activity matrix **U** can be decomposed as

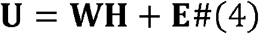

where **E** is the estimation error term matrix. The resulting matrix **W** and **H** are population version of primitive signals and synergy, respectively.

### Existing approaches

Next, here we compare the proposed method with two existing approaches to population mean estimation of motor modules: the module matching approach and cNMF approach. The major difference between the proposed method and the two existing approaches is that the population mean is estimated after first estimating each individual motor module. Denote by **X** the matrix whose elements are *μ*_*i,t,m*_ :

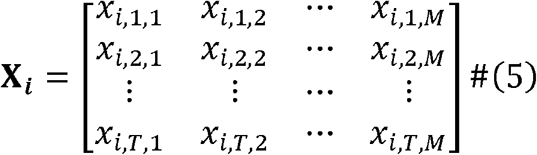

For the module matching approach, individual motor modules are first estimated:

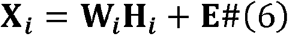

In contrast to the proposed method, the order of the synthesized variables (i.e., rows of **W** and columns of **H**) is arbitrary, so matching process is critical to make the modules correspond across subjects. One way to find the module of subject *j* that corresponds to the synergy *k* of subject *i* is to find a module of subject *j* that maximizes the correlation coefficient. That is,

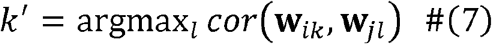

where **w**_*ik*_ and **w**_*jl*_ are column vector of matrix **W**_*i*_ and **W**_*j*_, respectively. To estimate the population mean of the motor module estimation, the mean of the matrix **W** or **H** is calculated for each subject after matching.

In this approach, the final **W** and **H** include mismatch errors due to the fact that the optimal number of modules (i.e., rows of **W** and columns of **H**) can be different across the subjects and there is no guarantee of a one-to-one correspondence between modules, and estimation errors resulting from local solutions due to initial value dependency of NMF algorithm^26,28^.

On the other hand, cNMF also performs individual module estimation, but it estimates individual motor modules by combining subjects as one pseudo-subject, applying NMF, and re-splitting the obtained results for each subject. To construct the input matrix, the individual muscle activity matrices **X**_**1**_,**X**_**2**,_ …, **X**_***n***_ are concatenated row by row

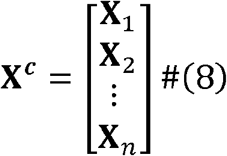

then, estimate the motor module:

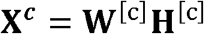

Splitting **W**^[*c*]^ into the original sample-wise blocks for 1,2, …, *n* corresponds to the primitive signal for each subject. Then, the population mean of the motor module estimation can be estimated by averaging these estimates. The cNMF approach assumes that the synergy **H**^[*c*]^ is common across samples^11^, which is implicitly considered equivalent to estimating the population mean **H** across subjects. Thus, cNMF can be otherwise described as a method of estimating **W**^[*c*]^ conditioned on the population mean **H**, which is expected to give results close to those of the proposed method.

The cNMF algorithm naturally results in larger matrices with increasing sample sizes, the use of NMF on these matrices can present challenges due to the higher number of parameters that must be estimated. First, the possibility of initial value-dependent local optimum solutions is further exacerbated. Also, the inclusion of some outliers can affect the process of sequential optimization, making it less robust to noise and increasing the likelihood of bias in the estimated results. It also increases the computational cost in terms of memory size and iteration to convergence.

### Evaluation with an actual data set

The proposed method was examined using the muscle activity during the gait cycle in seven young healthy subjects (**Figure 3A**). Based on the preprocessed muscle activities, the expected motor modules were estimated using three methods: the proposed method, module matching approach, and cNMF-based approach (**Figure 3B**). The number of motor modules was determined by the variance accounted for (VAF) (**Star Methods**). We selected 4 as the number of motor modules based on a VAF ≥ 99%.

**Figure 3.**
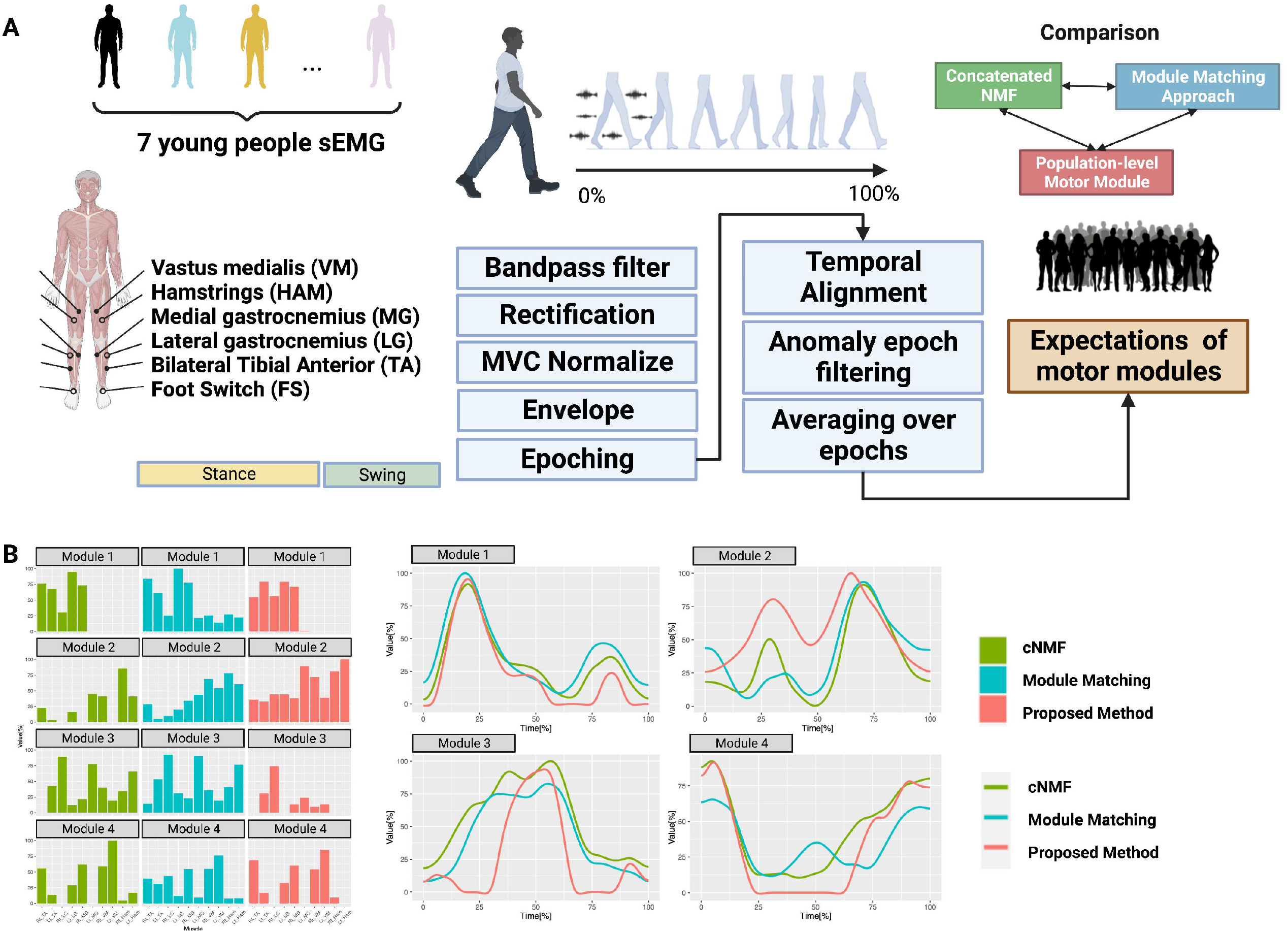
Actual data example. **A) Data overview and analysis design**. The Multichannel Electromyography (EMG) signals of 10 left and right muscles and foot pressure were measured in seven healthy young adults. The data were processed using an in-house analysis pipeline that implements standard muscle activity preprocessing. These preprocessed data were used to compare results from existing and proposed methods. **B) Comparison of estimated motor modules**. Four modules were identified based on variance accounted for. The left panel represents module composition **H**, the columns represent each method, and each row represents motor modules 1 through 4. Although all modules represent close trends between methods, there is variation in the estimates and different interpretations of the muscle groups that constitute the motor module. The right panel represents the primitive signal **W** corresponding to the four synergies, which also shows a similar trend between methods, but some of the modules show different timing of activation, suggesting that the clinical interpretation, such as the correspondence with the gait phase, can be different.

Although both methods showed similar trends for both **W** and **H**, inter-method variability in the estimates was evident (**Figure 3B**). For example, module 4 showed a marked difference in the timing of the increase in the primitive signal (**Figure 3B**). This could lead to substantially conflicting conclusions regarding the timing of CNS activity during gait cycles between the proposed and other methods. There was also significant variation between the methods in module estimation (**Figure 3B**), with the individual-based approach indicating that the entire muscle was involved in any module, whereas the cNMF-based approach and the proposed method suggested a sparse group structure. There were also significant differences between the cNMF-based approach and the proposed method, for example, in modules 3 and 4, indicating differences in the inference to the motor module during gait cycles (**Figure 3B**).

### Simulation model for evaluating methods

Next we evaluated the degree to which the expected value of the motor module could be reproduced for numerically generated population-level muscle activity data (**Figure 4A**). Equation (1) allows us to represent individual muscle activity patterns as samples from a probability distribution with a population mean function and variance. To obtain sample-wise muscle activity that follows a given probability distributions, we first decompose population expectation of muscle activity into several linear combinations based on basis functions (e.g., B-splines functions). The mean function *μ*_*m*_(*t*) can be decomposed as

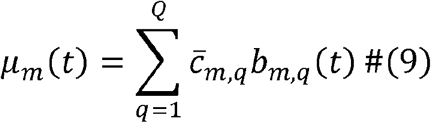

**Figure 4.**
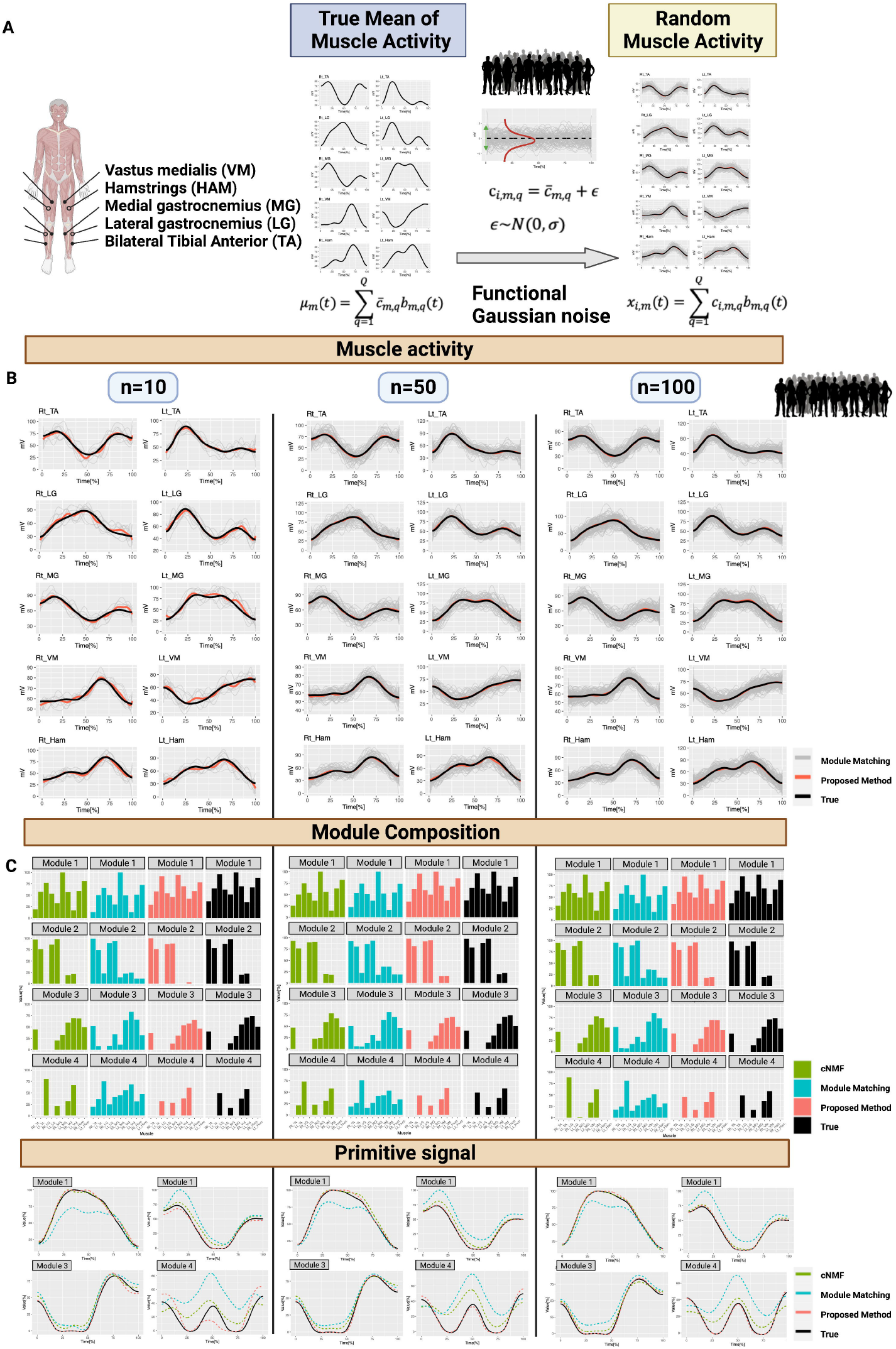
Simulation experiments of estimation bias. **A) Simulation model**. The model was constructed to generate Multichannel Electromyography (EMG) based on functional data representation. True muscle activity *μ*_*m*_ in skeletal muscle was approximated by linear combination by a small set of B-spline orthogonal basis functions *b*_*m,q*_ by regression splines with coefficients 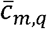. To mimic the simulation of real data, the mean muscle activity derived from the actual muscle activity of seven subjects was used. Assuming a normal distribution for the probabilistic distribution in the population, spline regression coefficients corresponding to individual subjects *c*_*i,m,q*_ were generated by adding noise which follows a multivariate normal distribution to 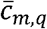, and by multiplying the B-spline basis *b*_*m,q*_ which was used for the true muscle activity approximation *μ*_*m*_. Resulting individual muscle activity profiles follow a normal distribution in the sense of functional data representation. **B) Generated individual muscle activity and its expectation**. The individual muscle activity profiles generated for n=10, 50, and 100 samples (gray lines) and their expected values (red) and true muscle activity (black) are shown. **C) Comparison of estimation bias of motor module expectation**. We estimated the expectation of motor modules and compared the estimation bias with each method. Here, we defined “true” expectation of motor module (black) as one estimated from true muscle activity *μ*_*m*_. The proposed method (red) gave an estimate close to the true motor module expectation (black) in both primitive signal and modules. On the other hand, exiting methods exhibited estimation bias in at least one of the modules.

A set of mutually orthogonal functions *b*_*m*.*q*_(*t*), are used to decompose the population mean function into *μ*_*m*_(*t*) functional linear combinations, each of which has weight coefficients 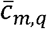. Here the coefficient vector 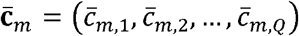 has sufficient information to reconstruct the population mean function under the basis function. In the FDA framework, the variation of muscle activity among subjects in a population can be described by variation with respect to this coefficient vector. In the numerical experiments, a normal distribution with mean average 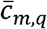and variance *σ* was assumed for individual muscle activity.

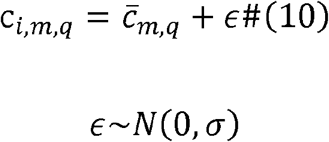

Substituting the sampled coefficient vectors back into equation (9) yields the individual muscle activities that follow the population distribution.

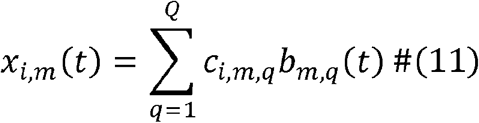

The FDA-based simulation model can be used to evaluate whether the expected values estimated from individual motor module estimates reproduce the expected values of the motor modules derived from the population mean of muscle activity. The residual sum of squares (RSS) of the true motor module was evaluated to assess the performance of each method. In other words, for the RSS of *k* −th module, we evaluated the

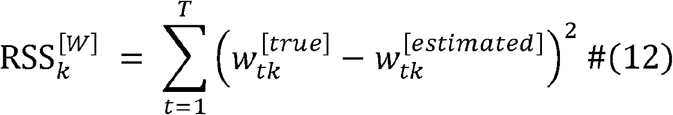

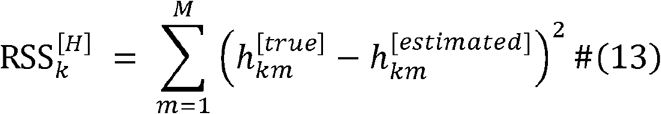

where 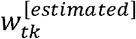 and 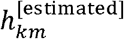 are estimated values for **W**^[*true*]^ and **H**^[*true*]^, respectively.

### The proposed model allows for control of estimation bias

We used a simulation model to assess each method (**Figure 4A**) with generated muscle activity for n=10, 50, and 100 (**Figure 4B**). While all methods captured the primitive signal’s trend, there was a bias in estimated motor modules based on individual and cNMF compared to the true motor module (**Figure 4C**). The individual approaches showed a tendency to overestimate the module weights i.e., averaged **H**_**i**_ in equation (6) over subjects after matching (**Figure 4C**). Our proposed method had accuracy close to that of cNMF, but with less estimation bias (**Figure 4D**). The fourth module showed an overestimation of the weight of the right leg TA (n=50) and underestimation of the left leg LG with cNMF (n=100) compared to the proposed method, with differences in the timing of peak signals (**Figure 4C, 4D**). We assessed the estimation bias of each method by calculating the RSS residuals for incremental changes in sample size. Our results showed that the proposed method converges to an estimation bias of zero as the sample size increases, in contrast to the other methods (**Figure 5B**). This suggests that existing methods lack asymptotic properties when estimating the population mean of motor modules.

**Figure 5.**
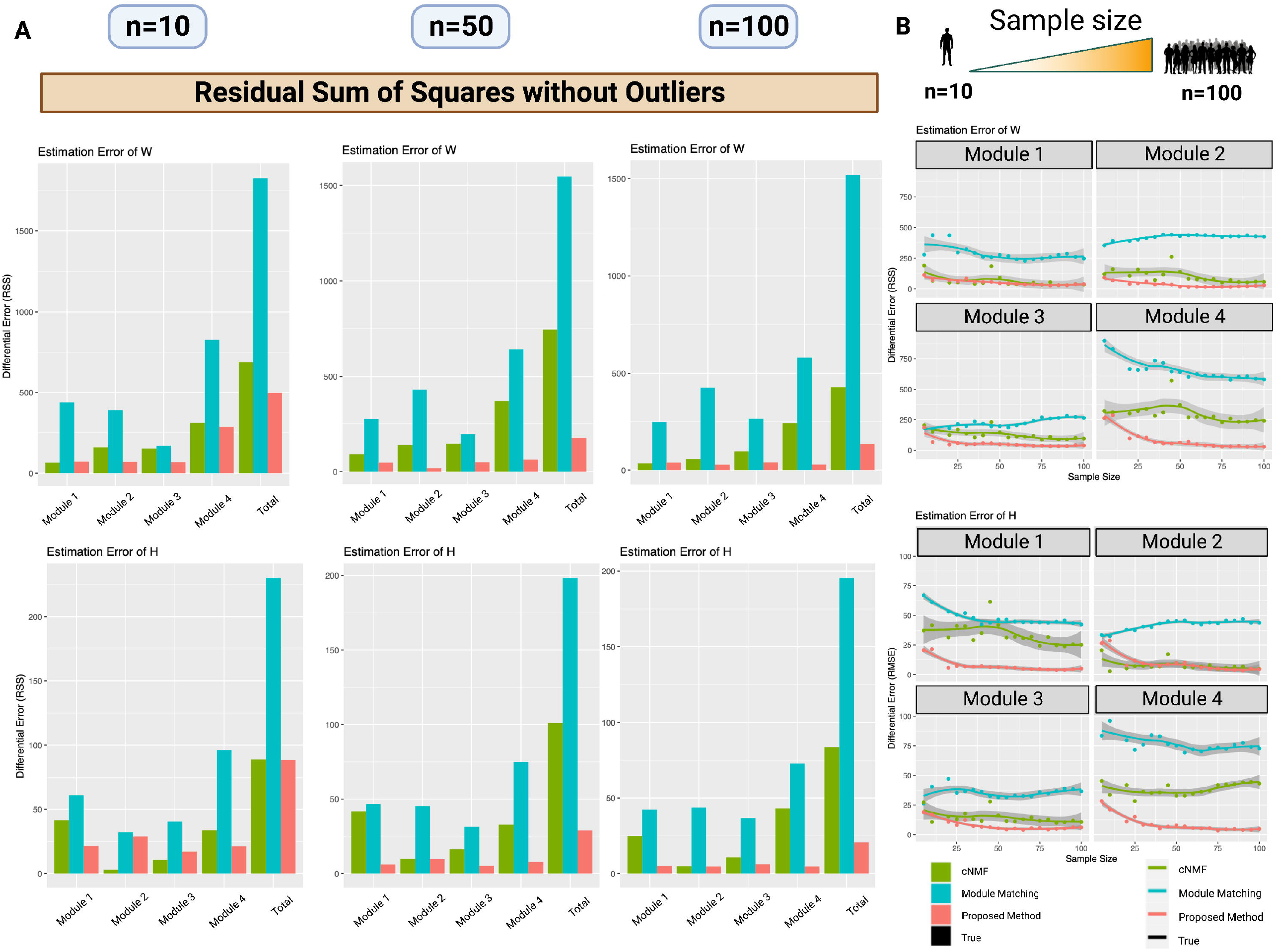
Comparison of residual sum of squares for estimation biases. **A) Accuracy of each method**. The residual sum of squares (RSS) calculated based on the results shown in Figure 4 are displayed for each motor module. The rightmost RSS in each panel shows the sum of the RSS over all modules. In the simulation, the proposed method (red) has the lowest RSS for all modules. **B) Asymptotics of RSS with sample size**. The RSS for each module is shown when the sample size is increased by 5 from 5 to 100 samples. With the nature of statistical error, RSS should asymptotically approach zero as the sample size increases. Only the proposed method (red) showed asymptotic behavior in the simulations.

### Evaluating noise robustness

To assess the impact of deviating values on estimation accuracy, we conducted a similar evaluation as in the previous section with 10% of the samples containing deviating signal values (**Figure 6A, 6B**). For small samples (n=10), all methods showed large estimation errors (**Figure 6C, 6D**). However, as the sample size increased (n=50, n=100), the proposed method significantly reduced the error compared to the other methods (**Figure 7A**). When we also examined the convergence of the estimation error with increasing sample size, the estimation bias of the proposed method converged to zero, while the other methods showed no tendency to converge (**Figure 7B**). Specifically, the estimation bias of cNMF increased significantly compared to the case without deviating values (**Figures 5A, 5B; Figures 7A, 7B**), indicating its sensitivity to such values.

**Figure 6.**
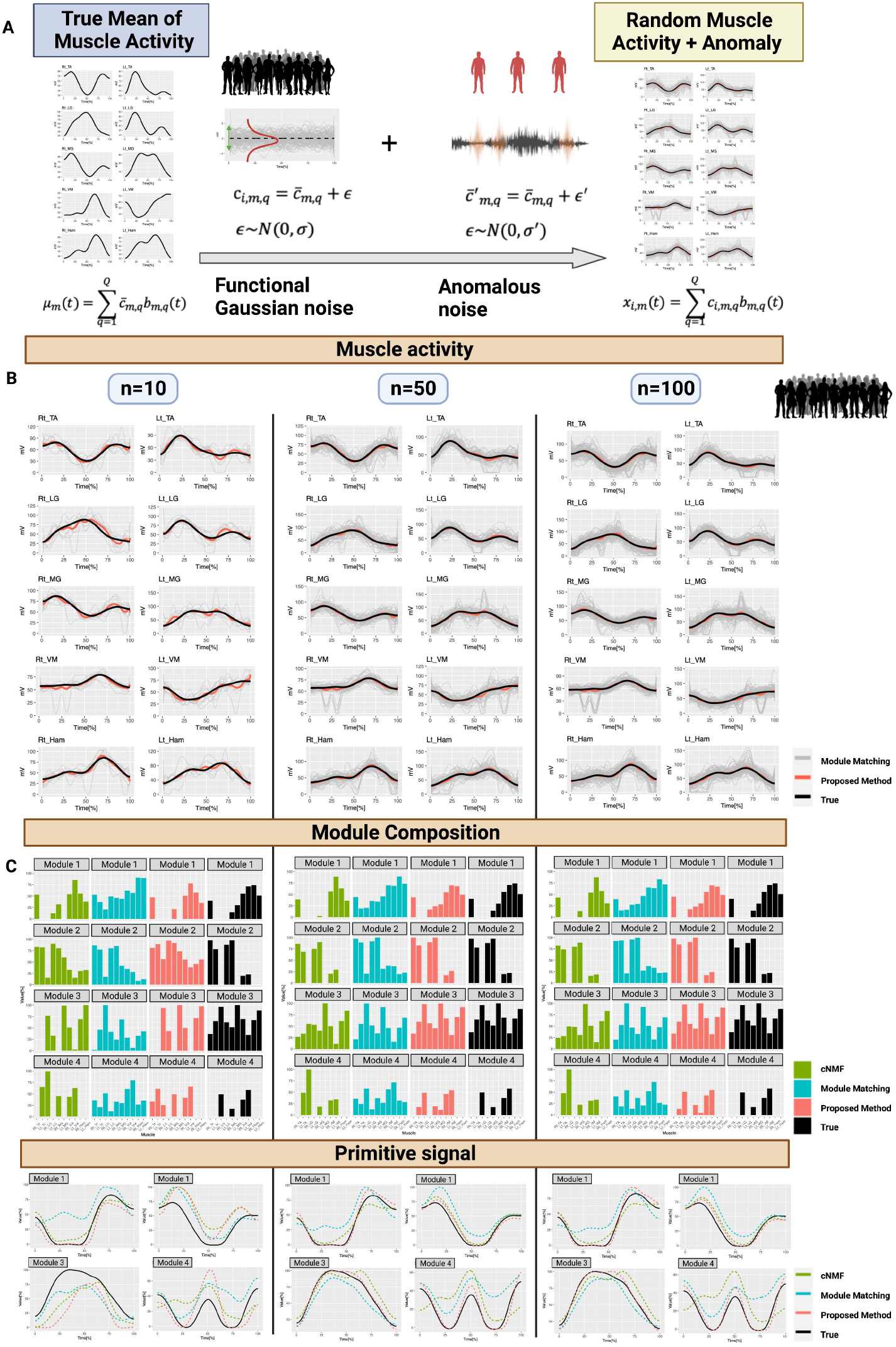
Simulation experiment of noise robustness. **A) Simulation model**. The simulation model shown in Figure 4 was modified to include some outliers. The muscle activity was generated by simulation assuming a population with 10% of outlier samples contaminated in addition to the normal sample. The generation of outlier samples was accomplished by adding errors with excess variance to a subset of the coefficients of the regression splines. The samples were then multiplied with the B-spline basis functions to obtain samples with distinct muscle activity profiles. **B) Generated individual muscle activity and its expectation**. The individual muscle activity profiles generated for n=10, 50, and 100 samples (gray lines) and their expected values (red) and true muscle activity (black) are shown. **C) Comparison of the estimation robustness of motor module expectation**. We estimated the expectation of the motor module and compared the robustness of estimation between methods. Under small-sample conditions, all methods showed estimation errors, but as the number of samples increased, the proposed method (red) gave estimates closer to the true motor module expectation.

**Figure 7.** Comparison of the residual sum of squares for robustness. **A) Robustness of each method**. The residual sum of squares (RSS) calculated based on the results shown in Figure 5 are displayed for each motor module. The rightmost RSS in each panel shows the sum of RSS over all modules. In the simulation, the proposed method (red) has the lowest RSS for all synergies, which indicates robustness to outliers. **B) Stability of RSS with sample size**. The RSS for each module is shown when the sample size is increased by 10 from 10 to 100 samples. It is expected that the effect of outliers will decrease as the sample size increases. However, only the proposed method (red) showed stability along with the sample size in this simulation.

## Discussion

We proposed an algorithm for estimating the motor module at the population level using Non-Negative Matrix Factorization (NMF) within the framework of Functional Data Analysis (FDA). Our method first estimates the population mean of muscle activity and then performs motor module estimation based on it. In contrast, the existing approach estimates the population mean after estimating the motor module for individual. The difference between the two approaches may seem simple, but our approach has been shown to significantly reduce estimation bias beyond a certain sample size and is also robust to noise.

Although our proposed algorithm showed convergence to the true motor module patterns with only 25 to 30 samples, the existing approaches failed as the sample size increased (**Figure 5B**). This was true even in the presence of samples that contained some deviant signal values (**Figure 6B**). Various factors can contribute to the failure of existing methods to converge. For example, the module-matching method can cause forced matching errors when there is no corresponding module, while cNMF may suffer from numerical instability or bias contamination due to collinearity or deviation values as the number of parameters to be optimized increases with the number of samples.

Our results indicate that our proposed algorithm outperforms existing methods in terms of noise robustness and convergence to true motor module estimates. However, we also found that the presented algorithm had a rather large estimation bias in small-sample conditions, and full convergence required larger samples (**Figure6B-D, Figure 7A, 7B**).

Therefore, while our proposed algorithm is an improvement over existing methods, its application may be limited in situations where the variability of the population is high and the sample size is very small. These results highlighted the need for statistical improvements in computational algorithms.

Alternative methods, such as principal component analysis (PCA) ^27,29,30^ and factor analysis (FA) ^31,32^, can also be utilized for motor module estimation, but in this study, we focused on NMF. One of the reasons for this choice is that NMF has been frequently employed and shown promise in estimation accuracy for various tasks, such as walking and running^13^. However, all these methods share common issues related to dimensionality reduction, such as the matching of synthetic variables’ order and sign, and potential contamination from heterogeneous observations, such as outliers^33,34^. Therefore, it is essential to further investigate and improve upon the accuracy and estimation methods for population-level motor module parameter estimation based on dimensionality reduction in the future direction.

This study has several limitations. Firstly, the validity of the estimation algorithm needs to be comprehensively evaluated for other commonly used movement tasks besides periodic walking. Secondly, the actual data analysis was performed using a small number of samples, and further evaluation with a larger clinical dataset is necessary. Thirdly, the presented algorithm assumes a homogeneous distribution of the latent population, and when multiple heterogeneous subpopulations are mixed, subpopulations must be identified and stratified. In this regard, future tasks include identifying subgroups based on dimension reduction^35^ and clustering^36,37^ and developing methods to test for heterogeneity based on functional data representation.

Characterizing motor modules at the population level is a crucial step towards translating experimental findings into clinical applications. For example, evaluation of intervention effects on motor function before and after intervention^38–49^, and large cross-sectional clinical studies^38,50,51^ need to capture effects as a population. The present study is the first to investigate the motor module estimation algorithm from a population-level perspective. Although the study has its limitations, such as the small sample size, it provides valuable insights into the estimation of motor modules at the population level. Continued development of the algorithms will fully realize their application to a broader range of movement tasks and clinical situations.

## Data Availability

All data produced in the present study are available upon reasonable request to the authors

## Acknowledgments

We would like to thank Editage (http://www.editage.jp) for editing and reviewing this manuscript for English language.

## Author Contributions

The study conceptualization was led by YM. Data analysis and interpretation were carried out by YM and IN. Algorithm development and evaluation were spearheaded by YM and KU. Manuscript drafting was a joint effort by YM and IN. All authors extensively reviewed, revised, and approved the final version of the manuscript for submission.

## STAR★Methods

### Resource availability

#### Materials availability

##### Data and code availability

The analysis code used in the paper can be accessed from the following repository URL (https://github.com/ymatts/RoMMS). The acquired data from this study are available from the lead contact upon reasonable request.

### Method details

#### Data acquisition

Wireless electromyograms (EMGs) (Trigno EMG sensors, DELSYS, Boston, MA, United States) were recorded from the bilateral tibial anterior (TA), soleus (SOL), medial gastrocnemius (MG), lateral gastrocnemius (LG), rectus femoris (RF), vastus medialis (VM), medial hamstring (MH), and lateral hamstring (LH) muscles, according to SENIAM recommendations. The skin was gently abraded and cleaned with alcohol before the EMG recording. The sensors were placed as far as possible each other anatomically to minimize the potential risk of crosstalk between the EMG recordings. Data were collected for two minutes at a self-selected speed and used to examine steady-state gait by removing the first 10 s of the gait. All the participants walked independently without walkers or crutches. The EMG signals were amplified (with a 909 gain preamplifier), band-pass filtered (10–450 Hz), and sampled at 1,000 Hz. Footswitches were attached to the heels to determine the time of foot contact.

### Preprocessing

The foot pressure signal values acquired from the foot sensors were used to identify the gait cycle. The time point at which the signal value increased from zero was defined as the heel contact. sEMG signals were processed for each muscle. They were first low-pass filtered at 30 Hz using a fourth-order Butterworth filter and then rectified. Maximum voluntary contraction (MVC) normalization was performed at the maximum observation as 100%. An epoch was created for each gait cycle and identified using a foot sensor. To identify and exclude abnormal cycles, each trial was projected onto a two-dimensional space using a robust principal component analysis^52–54^. Using these coordinate values, trials that deviated significantly from the projected space were defined as outliers and were excluded. The threshold settings for the anomaly values followed the default values in the R package rospca^55^. The variation in time between the gait cycles was corrected according to linear length normalization (LLN), with 0% for the first heel contact and 100% for the next heel contact. Finally, a 10-Hz envelope was derived, and the average was calculated to obtain the muscle activity.

#### Computational algorithm

Because the parameter of time *t* is observed as a discrete value in the real world, the muscle activity *x*_*i,m*_(*t*) is expressed as *x*_*i,t,m*_ using the discretized time *t* = 1,2, …, *T*. Equation (3) can be obtained using the time-discretized version of (4).

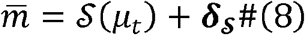

where and *μ*_*t*_ = (*μ*_*t*,1_, *μ*_*t*,2_, …, *μ*_*t,M*_) and

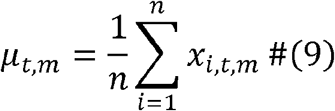

The explicit form of the motor module by NMF in equation (6) can be defined using muscle activity matrix **U**^[*T*×*K*]^ = {*μ*_*t,m*_; *t* = 1,2 …, *T, m* = 1, 2, …, *M*}. The primitive signals **W**^[*T*×*K*]^ and synergy **H**^[*K*×*M*]^ obtained by the NMF are derived as follows:

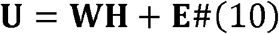

where **E** is the estimation error term with the *T*×*M* matrix.

#### Analysis pipeline

The workflow of the analysis is shown in **Figure 3A**. The first step was to perform standard signal processing on the raw sEMG signal for each subject, including bandpass filtering, rectification, MVC normalization, envelope smoothing, and epoching. Then, time alignment was performed, if necessary. Finally, the trial average was calculated for each muscle to characterize the muscle activity pattern for one trial per subject. We then estimated the expected mean of the muscle activity in the subject population according to equation (9) and applied the NMF to obtain an estimate of the population mean of the motor module shown in equation (10). For the number of modules, we used variance accounted for (VAF), defined below:

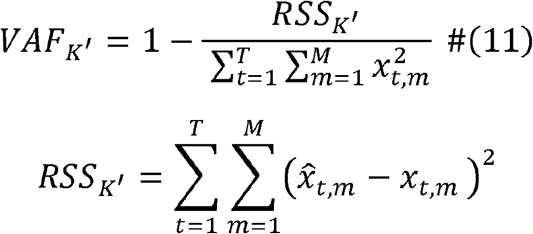

where 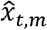 is reconstructed matrix of muscle activities using *K*^′^ synergy.

#### Simulation model

This section presents a simulation model for comparing the performance of each method. Two simulations are conducted. The first is the accuracy with which each approach can reproduce the “true” motor modules in the population (**Figure 3A**). Here, “true” refers to the motor modules “estimated from the true population mean of muscle activity.” Individual muscle activities were generated by adding normally distributed variations with a zero mean and a certain standard deviation to the predefined true population mean.

The second is the robustness of motor module estimation in the presence of outliers (**Figure 7A**). Assuming muscle activity from unrelated populations, the muscle activity distributed around different population means was prepared, and the muscle activity distributed around it was generated as an outlier sample. By mixing this with samples from the population of interest, we generated muscle activity that contained outliers and performed a simulation similar to the first to evaluate the reproducibility of the true motor modules.

The muscle activity patterns of each subject, which varied based on the population mean, were generated using the FDA framework (**Figure 3A**). Let the muscle activity in skeletal muscle *m* of subject *i* be the function value *x*_*i,m*_(*t*), and let the population mean be *μ*_*m*_(*t*) and the population variance be 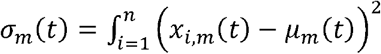. Functional observations can be expressed using several orthogonal basis functions *b*_*m,q*_(*t*);*q* = 1,2, …, *Q*.

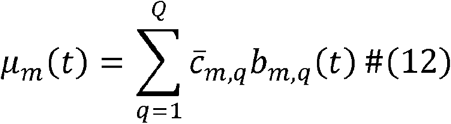

This simulation used a B-spline basis, which is a typical orthogonal basis function in the FDA. The coefficients 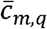 were estimated based on a regression spline and the number of basis functions were determined by cross-validation. To mimic the actual muscle activity pattern, we derive the mean function *μ*_*m*_(*t*), which we would like to estimate, from a real dataset. The average of the preprocessed muscle activity of the seven young healthy subjects was calculated, and this was set as *μ*_*m*_(*t*).

For the coefficient vector 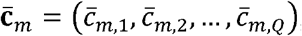, a subject-specific coefficient vector ***c***_*i,m*_ = (*c*_*i,m*,1_, *c*_*i,m*,2_, …, *c*_*i,m,Q*_) was generated by adding an independently generated noise normal distribution, which is considered biological variability (**Figure 4A**). That is, for *q* = 1,2, …, *Q*.

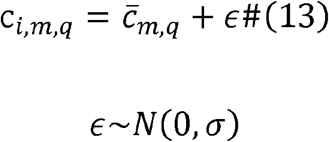

In this simulation, we set *σ* = 1 assuming that the subject population follows a standard normal distribution. Finally, the orthogonal basis functions were multiplied again to generate the functional observations for each subject.

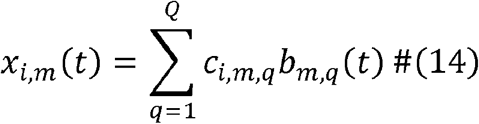

We compared the algorithms for estimating the population mean of the motor modules using muscle activity **X**_***i***_ = {*x*_*t,m*_} consisting of *x*_*i,t,m*_ discretized by in equation (14).

For another simulation of the effects of outliers, heterogeneous samples were mixed (**Figure 6A**). The muscle activity patterns of these samples were generated using equation (13). That is, assume a different population of no interest with population mean *μ*_*m*_(*t*) ;

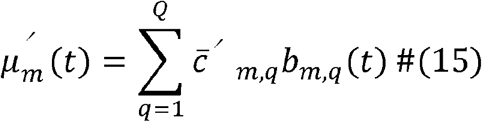

The coefficients 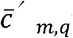 were randomly selected from the coefficients 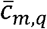 of the population mean of interest in equation (9), to which was added a noise of normal distribution with large standard deviation *σ*^*′*^ (> *σ*) (**Figure 6A**);

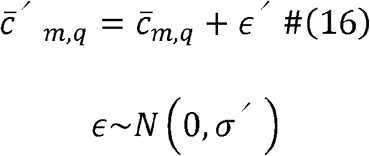

In this simulation on the effect of outlier two coefficients *r*_1_, *r*_2_ (≤ *Q*)) were randomly selected from *q* = 1,2, …, *Q* and equation (13) was applied and obtained outlier samples;

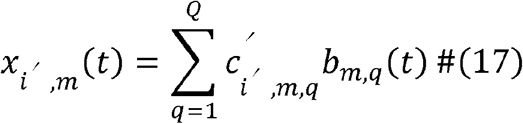

The percentage of outlier samples was simulated as 10% of the total sample size, *N*.

#### Evaluation of methods

In this section, we evaluate the performance of these methods using a simulation dataset. We defined the motor module derived on the basis of equation (12) as the “true” motor module.

Let **W**^[*true*]^ and **H**^[*true*]^ denote the true motor module estimates obtained by using the NMF algorithm. Each matrix element is denoted as 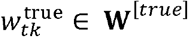 and 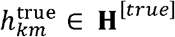.

Here, the primitive signal is represented as **W**^[*true*]^ and the weight of each muscle, that is, the module, is denoted by **H**^[*true*]^.

The residual sum of squares (RSS) of the true motor module was evaluated to assess the performance of each method. In other words, for the RSS of *k* −th synergy, we evaluated the

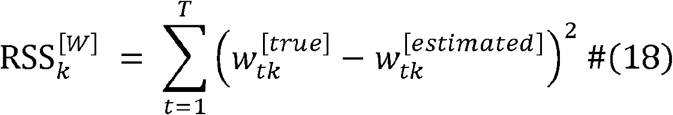

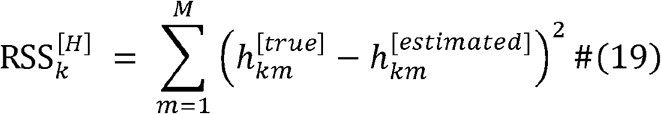

where 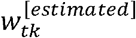 and 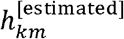 are estimated values for **W**^[*true*]^ and **H**^[*true*]^, respectively.

Note that we normalized the values 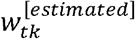 and 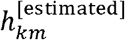 for each method because the scale of the estimated values differs from method to method. Normalization was performed for each estimated motor module by each method; each element was divided by the value of maximum value in the estimated primitive signal **W**^′ [*estiamted*]^and synergy ′ **H**^′ [*estiamted*]^ and multiplied by 100. This was also true for ′ **W**^[*true*]^ and ′ **H**^[*true*]^. After normalization, these estimates were used to calculate the RSS using equations (18) and (19).

